# A Systematic Review and Meta-analysis of Polycystic Ovary Syndrome and Mental Health among Black Asian Minority Ethnic populations

**DOI:** 10.1101/2022.03.05.22271948

**Authors:** Gayathri Delanerolle, Salma Ayis, Vanya Barzilova, Peter Phiri, Yutian Zeng, Sandali Ranaweera, Ashish Shetty, Nyla Haque, Debasish Kar, Kingshuk Majumder, Shanaya Rathod, Vanessa Raymont, Jian Qing Shi, Dharani K. Hapangama

## Abstract

**Background:** Polycystic Ovary Syndrome (PCOS) is a chronic and a common gynaecological condition impacting women of reproductive age. Women with PCOS have hormonal, ovulatory and metabolic dysfunction resulting in multiple symptoms. The correlation between hormonal disbalance and the impact on women’s mental health (MH) has been researched for decades. However, the prevalence among different ethnicities has not been fully evaluated.

**Methods:** A systematic methodology was developed, and a protocol was published in PROSPERO (CRD42020210863) and a systematic review of publications between 1^st^ January 1990-30^th^ January 2021 was conducted. Multiple electronic databases were explored using keywords and MeSH terms. The finalised dataset was analysed using statistical methods such as random-effect models, subgroup analysis and sensitivity analysis.

**Findings:** We included 30 studies reporting on 3,944 PCOS women. Majority of studies addressed depression anxiety, and common mental health. Studies had fair to poor methodological quality and includes observational studies and Randomised Clinical Trials (RCTs). Overall, 17% (95% CI: 7% to 29%) of women with PCOS have clinical diagnosis of major or severe depression; 33% (95% CI: 26% to 40%) have elevated depressive symptoms or a clinical diagnosis of depression; 41% (95% CI: 28% to 54%) report anxiety symptoms, and 31% (95% CI: 15% to 51%) have a form a common mental health or are takingpsychiatric medication for anxiety and / or depression. The use of various tools to assess mental health symptoms was among the reasons for the substantial heterogeneity across studies.

**Interpretation:** PCOS is associated with an increased risk of mental health disorders including depressive s, anxiety, and other mental health disorders. While BAME populations account for about 20% of most of the samples studied, stratification by ethnicity was rarely attempted which made it difficult to elucidate the MH impact of PCOS on different communities.

**Funding:** Not applicable

**PROSPERO registration number:** CRD42020210863

## Introduction

Polycystic ovary syndrome (PCOS) is the most common endocrine disorder in women of reproductive age with significant reproductive, metabolic, and psychological implications. An estimated 8-13% of women of reproductive age are affected,^1–5^ with a widely quoted similar prevalence between different ethnicities.^6^ The Rotterdam criteria,^5,7^ which incorporates a combination of signs and symptoms of androgen excess, ovarian dysfunction and polycystic ovarian morphology on ultrasound is the globally accepted diagnostic criteria for PCOS. The phenotype between different ethnicities, can vary markedly^5,8,9^ with PCOS sufferers from Hispanic, East Asian, South Asian and Middle Eastern backgrounds displaying higher prevalence of symptoms associated with hyperandrogenism^8,9^ than their White counterparts. The precise aetiology of the syndrome is unclear, although evidence suggests a genetic predisposition, and a role of environmental variables including diet and lifestyle factors.^10,11^ The pathophysiology involves a dysfunctional hypothalamic-pituitary-ovarian axis, with inappropriate gonadotropin-releasing hormone (GnRH) pulsatility and increased pituitary secretion of luteinizing hormone (LH).^12^ Those affected with PCOS present with a wide spectrum of clinical symptoms and features, which can be reproductive (oligomenorrhoea, hirsutism, subfertility), metabolic^13,14^ (insulin resistance, type 2 diabetes, metabolic syndrome), and psychological^15,16^ (depression, anxiety and low quality of life).

PCOS, due to its diversity in clinical presentation, remains undiagnosed in up to 70% of women,^3^ this may lead to various clinical sequelae that impact and complicate women’s long-term health. For example women with PCOS have an increased risk of consequential complications such as type 2 diabetes mellitus, cardiovascular disease, endometrial cancer, pregnancy complications and psychological conditions.^17–20^

One area lacking in PCOS research are the MH correlates of the syndrome. The limited evidence available suggests that women with PCOS have higher rates of depression, anxiety, binge eating disorders, bipolar disorder and other psychological symptoms,^21–23^ most likely due to the androgenic, reproductive, and metabolic disorders or due to the associated symptoms of the disorder.^24,25^ Current treatment for PCOS focuses on physical symptom control and reducing complication rates.^26^ At a global level, currently, there is limited recognition of mental health (MH) symptoms in women with PCOS.^5^ However, background MH sequelae may significantly impact treatment adherence and patient quality of life.^5^

MH disorders account for 14% of global burden of disease.^27^ Yet significant disparities persist in providing access to MH services for Black, Asian and Minority Ethnic (BAME) populations.^28^ This is further exacerbated by the underreporting of MH symptoms in BAME individuals who are less likely to contact their general practitioner regarding their MH compared to their White British counterparts.^28^ This can be explained by a combination of personal and environmental factors, including inability to recognise MH symptoms, cultural identity and stigma, language barrier and poor communication with healthcare professionals.^28^ Therefore, the prevalence of MH disturbances in BAME women with PCOS remains unclear. The MH symptom prevalence rates may differ significantly across races and ethnicities for women with PCOS in the form of either symptoms and/or psychiatric disorders. Understanding the multifactorial and heterogenic nature of PCOS presentation in BAME women is crucial to improve clinical and patient reported outcomes.

An assessment of the existing evidence is vital to generate comprehensive knowledge and clinical practice improvements that are suitable for BAME populations. Therefore, a systematic review was designed and developed within this study.

## Methods

A systematic methodology was developed and peer reviewed prior to publication in PROSPERO in accordance with Preferred Reporting Items for Systematic Reviews and Meta-Analyses (PRISMA).

### Research question

To assess the prevalence of the disease sequalae between PCOS and mental health symptoms and disorders among BAME populations.

### Search Strategy

The search strategy involved multiple databases of EMBASE, PubMed, PsycINFO, Science direct and Web of Science from inception to the 30^th^ of May 2021. The searches were conducted through references initially followed by the review al their abstracts and tracking of citations.

### Eligibility criteria

All peer review publications in English were included to this study. Key words and MeSH terms included *depression, polycystic ovary syndrome, PCOS, bipolar, dysthymia, major depressive disorder, anxiety, psychosis, somatic symptoms, psychotic disorders, trauma, stress, post traumatic stress (PTSD), obsessive-compulsive spectrum disorders, ethnic minorities, BAME, BME and eating disorders*. Studies that included a psychiatric diagnosis or MH symptomatologies based on a clinically structured diagnostic interview and/or clinically accepted screening tools were eligible for this study. Meeting abstracts were excluded if there was no full paper available.

### Data extraction and analysis

Predefined clinical variables were used as part of the data extraction of rates of depression, anxiety, stress, schizophrenia, psychosis and PTSD within the search strategy. An evidence synthesis methods protocol was developed and has been shown as supplementary document 1. The data extraction process was completed as per the PRISMA guidelines whilst, the refinement protocol has been shown in the supplementary document 1. The extracted data was reviewed using Endnote and Microsoft Excel by 3 reviewers prior to the statistical analysis. All studies collated were categorised based on MH symptoms versus psychiatric disorders and, where possible synthesised based on prevalence and 95% confidence intervals. Prevalence tables were developed to indicate any subgroup categories including geographical location, race, heterogeneity, age, obesity scores and ethnicity. Systematically included studies that did not meet the meta-analysis criteria due to insufficient statistical data or poor quality were narratively synthesised and analysed based on the disease, family, clinician and patient perspective. The narration would include the reporting of common themes including potential barriers to identify and report any themes or sub-themes that may be present in clinical guidelines.

### Outcomes

The primary outcome was to assess the odds ratios and p-values associated with a possible mental health symptomatologies and/or psychiatric disorders among PCOS BAME populations.

### Statistical analysis plan

Depression was measured by a range of scales and several tools including the Diagnostic and Statistical Manual of Mental Disorders (DSM)^29^; the Patient Health Questionnaire (PHQ)^30^; the Hospital Anxiety and Depression (HAD)^31^; Beck Depression Inventory (BDI)^32^; Montgomery Åsberg Depression Rating Scale (MADRS-S)^33^; Center for Epidemiologic Studies Depression (CES-D)^34^; Reynolds Adolescent Depression Scale, 2nd Edition(RADS-2)^35^; Quick Inventory Depressive Symptomatology (QIDS)^36^; Self-Reported Questionnaire Scale-20 (SRQ-20)^37^; the Generalized Anxiety Disorder (GAD)^38^; the International Neuropsychiatric Interview (MINI),^39^ use of antidepressants or use of anti-anxiety medication; a diagnosis by a MH professional, and self-reported history of depression.

The types of mental illness considered by most studies were anxiety, depression, both anxiety and depression, and generalised mental wellbeing. The prevalence of these was synthesised using a meta-analysis.

For the purpose of this study, depression was defined as a major depression diagnosis or elevated depressive symptoms, as reported within the systematically included studies. The diagnosis of major depressive disorder (MDD) was based on clinical review which included suicidal ideations, or provided a definition of MDD based on thresholds of tools used, for example, BDI ≥ 17, CES-D ≥ 24.^40,41^ Elevated depression included clinical depression, antidepressants use, and / or a threshold for mild to moderate depression based on the scale used for assessment, for example BDI ≥ 11, HAD-D ≥ 8, and CES-D ≥ 16. For anxiety, a combinational approach was used where the use of the term anxiety or use of anxiety medications were pooled together, due to overlapping definitions, and lack of clear cuts for severity levels. Generalised mental wellbeing on the other hand was defined, as having both depression and anxiety, use of medication for psychiatric disorders as reported within the studies, self-reported MH symptoms, or reported under the term “common mental health” (CMH) by a study authors based on scales such as SF-36.^42–44^

A Random-effects models were used to calculate overall summary estimates of anxiety, depression, and CMH in PCOS patients.^45^ The routine “metaprop” of the software STATA’s (V.16) which provides pooled summary of proportions and 95% confidence intervals based on Newcombe exact binomial estimation was used.^46,47^ The routine also estimates the I-squared statistics which describes the percentage of variation across studies that is due to heterogeneity rather than chance. (Higgins et al. 2003)^48^

All studies systematically included demonstrate a range of study designs including of observational and randomised clinical trials (RCTs). Where comparisons were made between PCOS and non-CPOS groups in a study, only the PCOS groups were included. At situations where PCOS population were stratified by certain criterion, for example body mass Index (BMI) or age, the strata were combined where the sample size was smaller than 30 for each group, and an overall prevalence was calculated. For larger groups, each was treated a separate group. Sensitivity analyses in addition, were used to highlight differences across strata where a criterion proven to have an impact on heterogeneity.

Publication bias was assessed using funnel plots and Egger test, for symmetry and small study effects where the number of studies exceeded the recommended 10.^49,50^

### Risk of Bias (Quality assessment)

The Newcastle-Ottawa-Scale (NOS) was used to assess the risk of bias (RoB) for all systematically included studies as demonstrated within the RoB table.

### Sensitivity analysis

Sensitivity analysis was used to explore causes of heterogeneity. BMI, and tool of assessment of assessments were used for stratification.

## Results

Figure 1 displays the estimated prevalence and 95% confidence intervals (CI) for the mental health conditions classified as: (a) Major depressive disorder, (b) Elevated existing depression, (c) Anxiety and (d) Common mental health.

**Figure 1.**
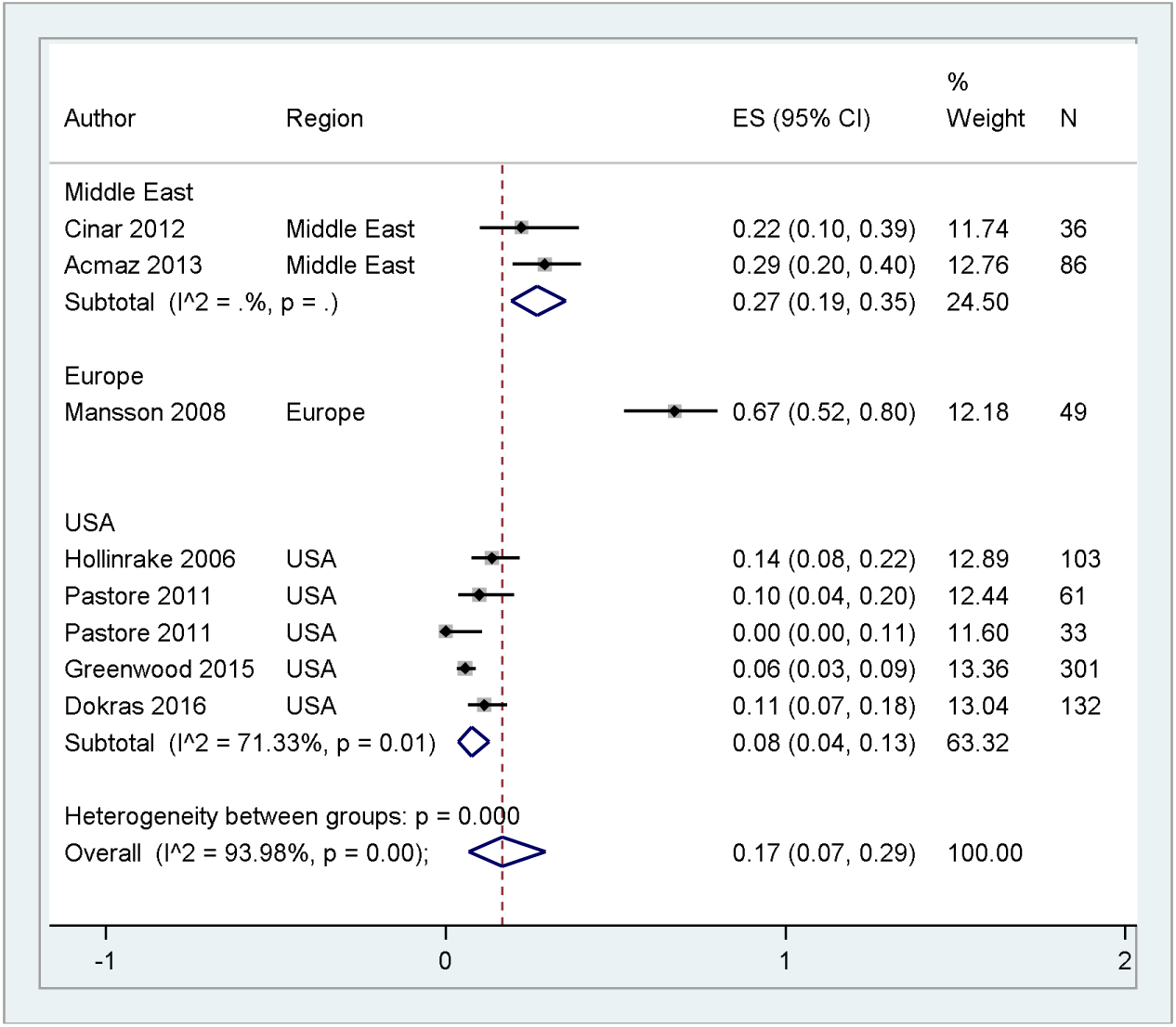
The Prevalence of Mental Health Indicators in Women with PCOS (a) Major depression by region

**(b).**
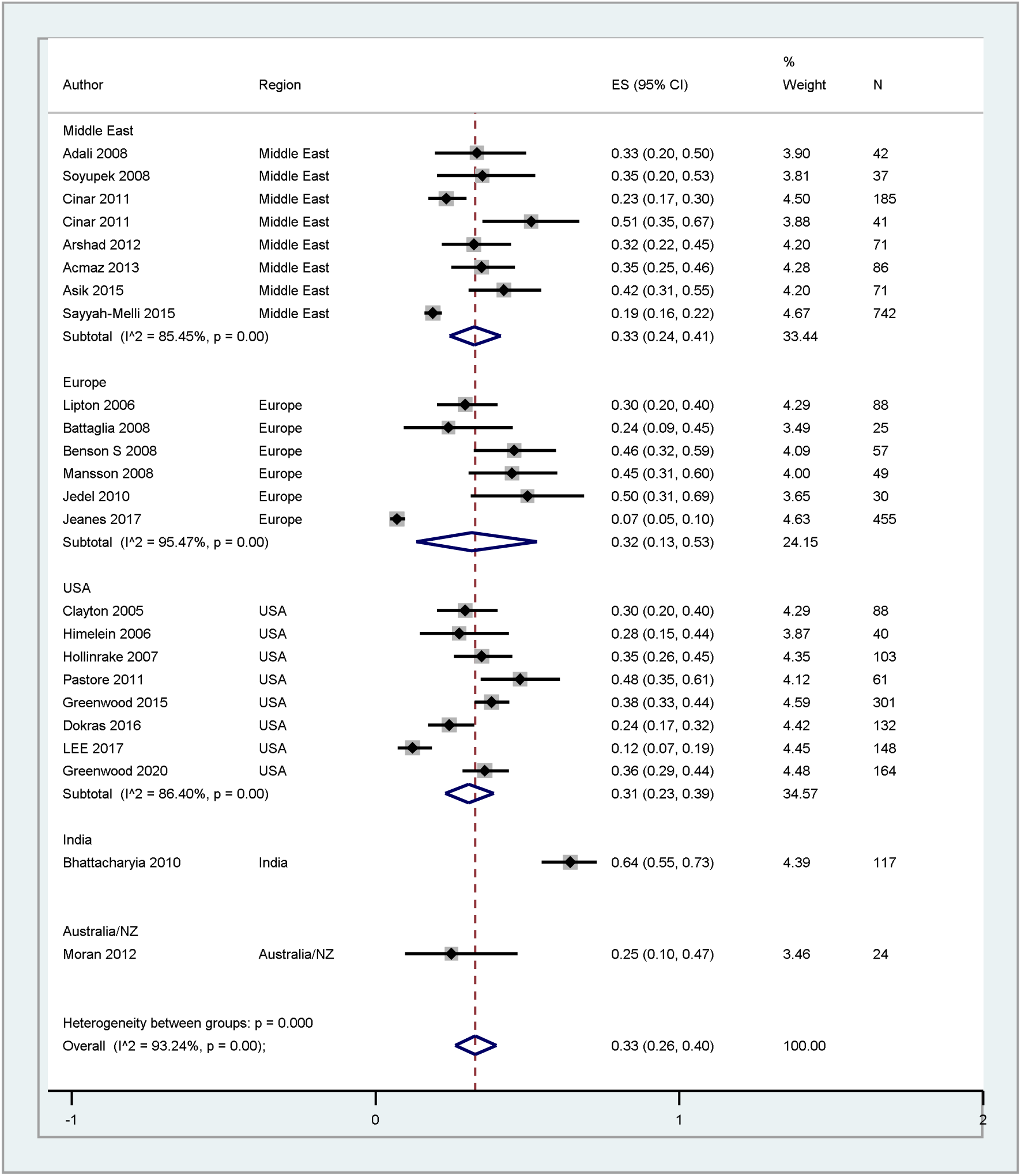
Elevated depression by region

Overall, major depressive disorder affected 17% of PCOS patients, and vary between 0% to 67%. All studies except two have estimated a prevalence ≥ 10%. The higheset prevalence was reported in a Swedish study, followed by two Middle Eastern (ME) studies, with a prevalnce of 67%, 29%, and 22% for the three studies respectively.^51–53^ Substantial heterogenity, I^2^ of 93.98% was reported between regions, also considerably high within USA studies, I^2^ of 71.33% but the two ME studies were homogeneous, Figure 1 (a).

Depression was summarised from 24 studies, including 3157 women with PCOS. The estimated average was 33% (95% CI: 26% to 40%). Where the estimated prevalance was startified by region, fairly similar estimates on average, were noted across regions, with India being the only exception where the the highest prevalence of 64% (95% CI: 55% to 73%) was reported. Substantial heterogeneity observed within regions and between regions. The overall hetrogenity was 93.24%.

For anxiety, Figure 1 (C), the highest prevalnce of 75% was seen in a USA study, with 78% white and 22% non-white, mean age 33 years (SD: 7.5), followed by a prevalance of 74% in a European study with 80% white, 18% Asian, age 33 years (7.4), while the two studies didn’t report BMI.^54,55^ Middle East, and Australia/ NZ follow with an estimated averages of 42% and 38% respectively, and the overall estimated prevalnce was 41% (95% CI: 28% to 54%).

**(c).**
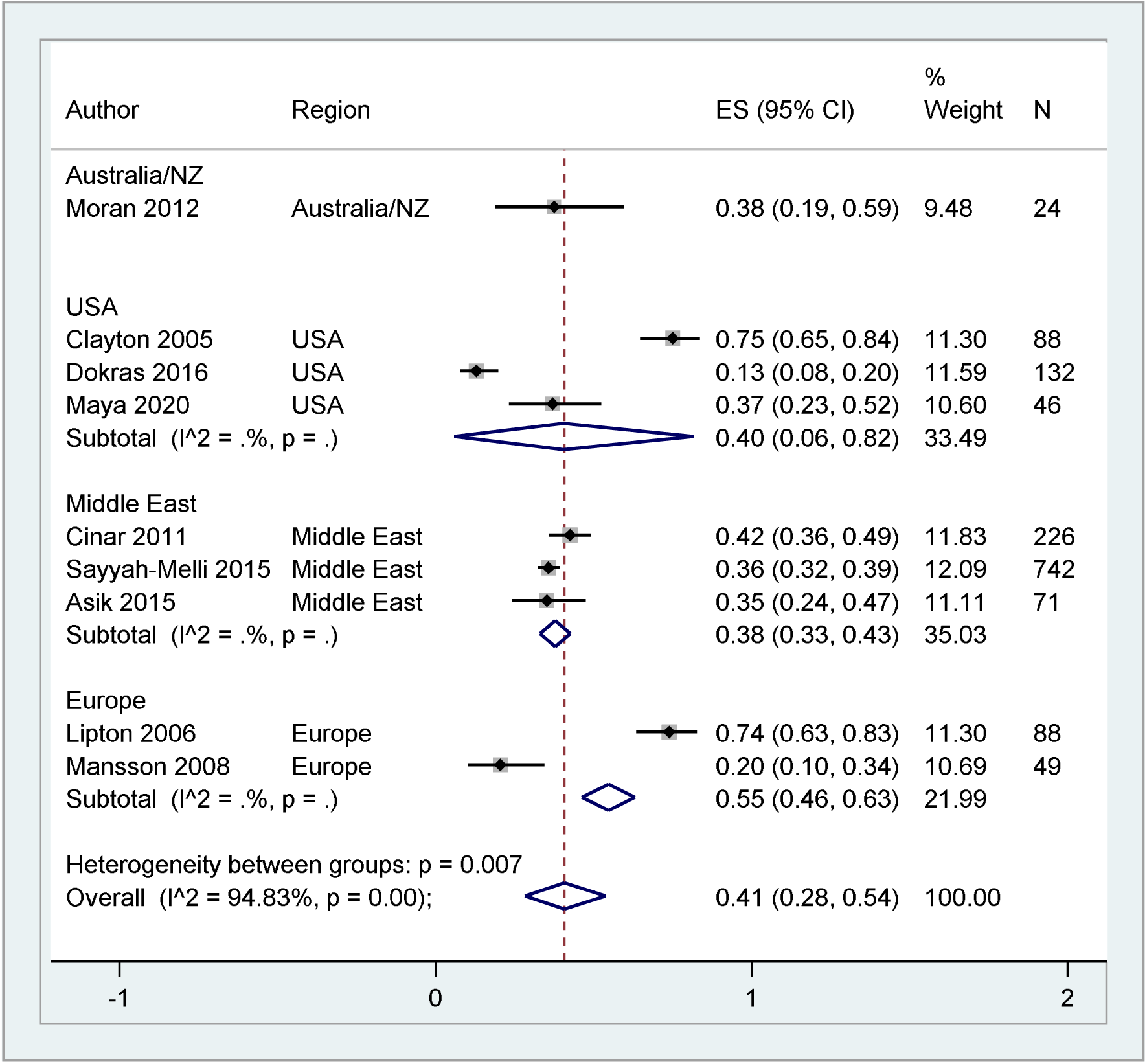
Prevalence of anxiety by region

Figure 1 (d), common mental health overall prevalence was 31% (95% CI: 15% to 51%). Substantial heterogeneity shown by USA studies, only one study from ME, and one from Brazil were included. A substantial lack of consistency across studies was reflected by a heterogeneity I^2^ of 97.29%.

**(d).**
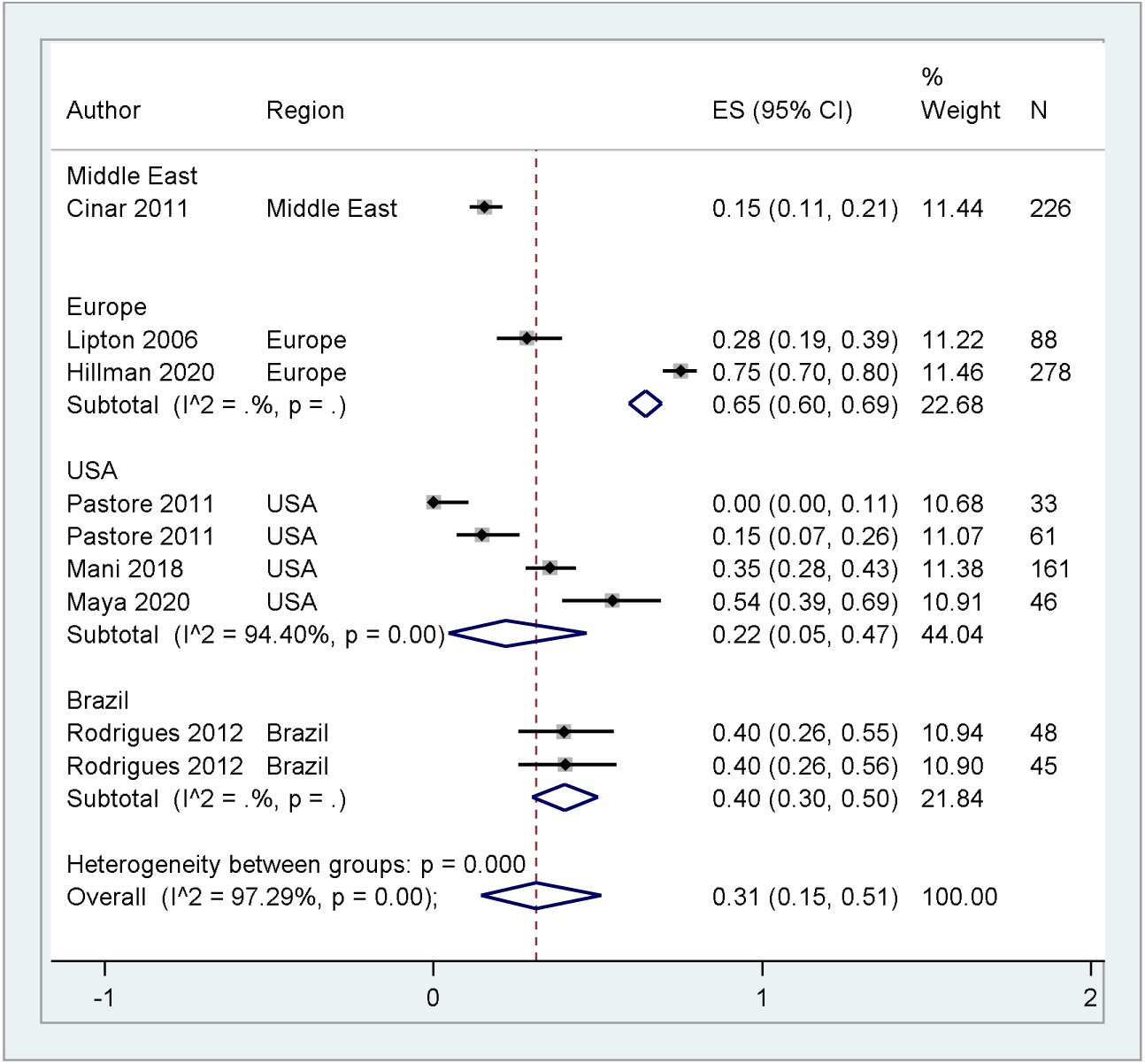
Common mental health by region

**FIGURE 2.**
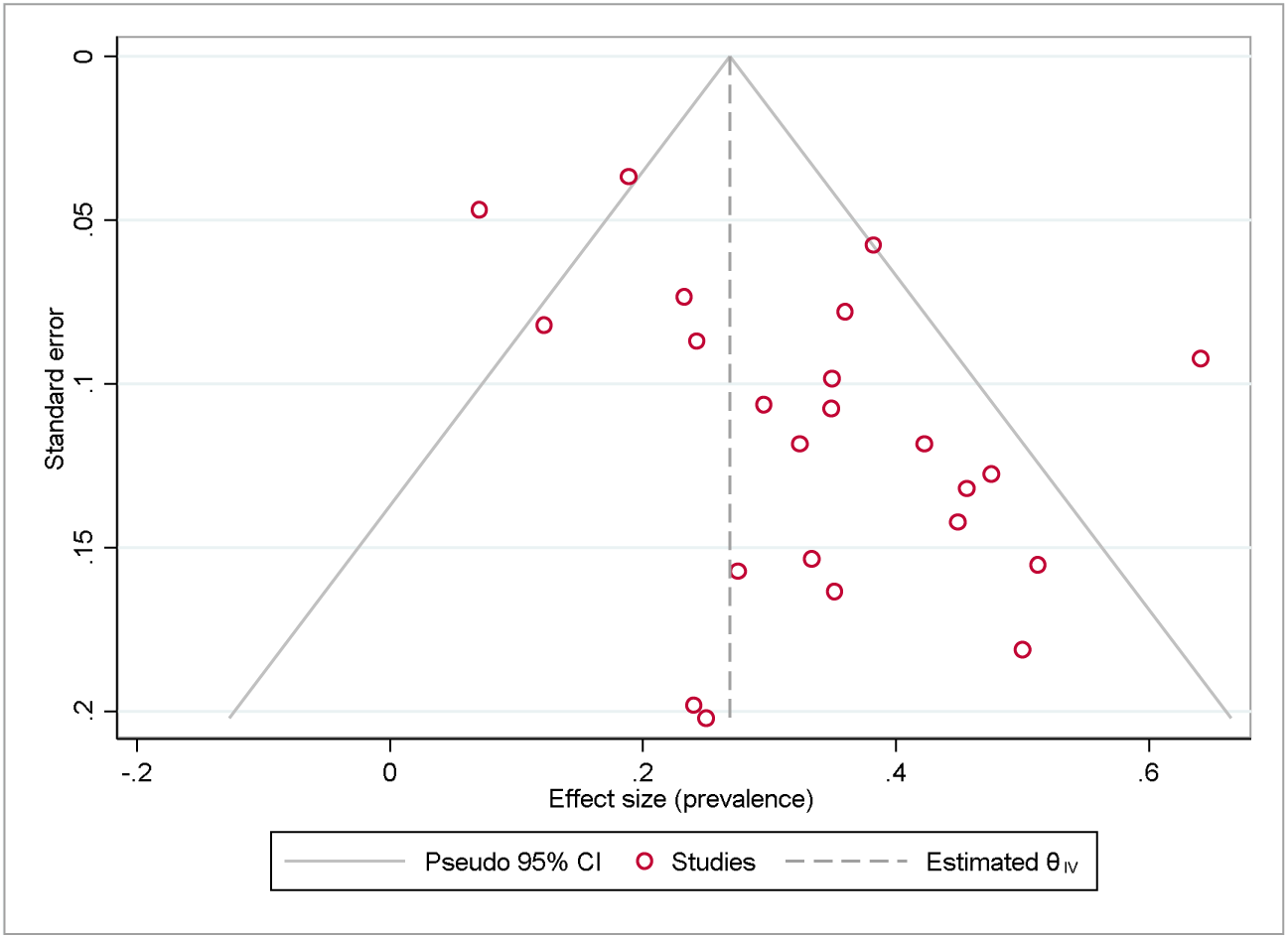
(a) Funnel Plot (Prevalence of elevated depression) Note. The figure visually supports an asymmetrical shaped funnel plot. The no small study effect null hypothesis (H0: no small-study effect) may therefore be rejected. Using Egger test of H0: no small-study effects, p value = 0.004.

**(b).**
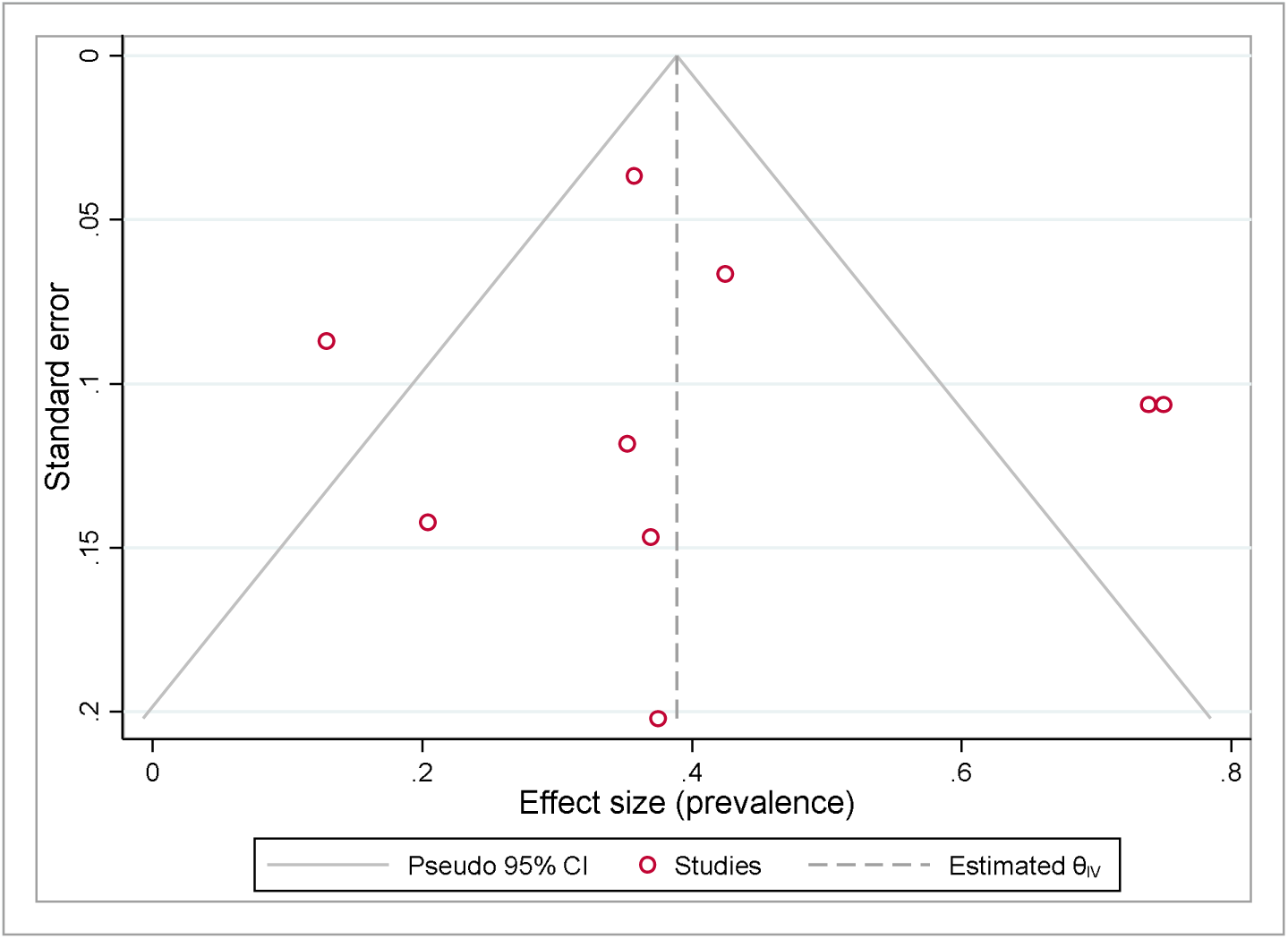
Funnel Plot (Prevalence of anxiety) Note. The figure visually supports the symmetry of the effect size distribution. The correlation between effect size and the corresponding standard error was not strong, Egger’s test p value = 0.65 supporting no small-study effect (symmetry) hypothesis.

## Discussion

The meta-analysis identified 11 studies which reported a 20% prevalence of common MH symptoms of anxiety and depression primarily. However, there was substantial heterogeneity across all studies which is reflected by I^2^ of 97.29%. Sources contributing to this heterogeneity may include differences in age range, body mass index (BMI), region, study design (including psychiatric assessments used), sample size and methodology, population ethnicity and other characteristics. The narrative synthesis identified several key themes including symptoms of depression and anxiety, as well as diagnosed conditions of eating disorders, bipolar disorder, PTSD and psychotic disorders as demonstrated in Table x.

Despite being the most common reproductive endocrine disorder that spans through the entire reproductive life of a woman, PCOS is often associated with a significant negative patient experience, including delayed diagnosis, that could attribute to exacerbation of symptoms.^56–58^ Due to the chronic nature of the disease, challenges with persistent treatment adherence is a common issue, but lifestyle changes can be useful to decrease the long-term consequences of the disease. However, once diagnosed, only 19% of women report being aware of all conditions associated with PCOS following discussions with their general practitioner (GP).^59^ The typical symptoms of PCOS such as disturbed menstrual cycles, hirsutism, male-pattern baldness, acne, obesity, and infertility^12,60–63^ could have a further negative impact on patient self-esteem leading to psychological burden. However, narrative accounts point to considerable emphasis on a few symptoms such as infertility by the clinicians, rather than taking a holistic view on all important features of the condition,^59^ such as the long term physical and mental health sequalae, and the effect of diagnosis and treatments on quality of life of the woman. PCOS is also associated with a higher rate of psychiatric disorders.^56^ In keeping with this, our meta-analysis reported 31% prevalence of common MH symptoms and diagnoses in women with PCOS. Similarly, Hillman et al. found an overwhelming 74.9% of women diagnosed with PCOS in their study, to have reported that the condition has adversely affected their MH.^59^ However, despite the considerable negative effect on psychological wellbeing, as many as 34.9% of women in the study chose not to discuss associated MH symptoms with their GP.^59^ An important factor contributing to this may be the ethnic background of the patient. Hillman et al. report that caucasian women in UK diagnosed with PCOS were more likely to discuss their MH with their GP when compared with asian women in UK with PCOS (odds ratio [OR] = 1.66; 95% confidence interval [CI] = 1.04 to 2.63).^59^ Clinicians and healthcare providers should better evaluate the needs of BAME communities. This would allow personalised care to be delivered in a culturally sensitive way, which may improve overall patient and clinical outcomes.

Around 1 in 6 adults in England meet the diagnostic criteria for common MH disorders.^64^ BAME ethnic groups are more likely to be detained under the Mental Health Act compared to their White British counterparts, and this can be explained by higher rates of mental illness observed and poorer levels of support.^65^ Several studies have explored beliefs about mental illness within BAME communities, and the barriers faced when accessing care. Factors which account for poor presentation rates with mental illness amongst the BAME community include associated stigma, including those with PCOS, poor English language skills, previous negative experiences with healthcare professionals, and being aware of available services.^66^ The MH of patients with PCOS may be severely impacted by these issues and reluctance to seek help may further exacerbate pre-existing psychiatric conditions, or limit the recognition of new MH symptoms. Therefore, a thorough assessment of MH illness risk is particularly important in BAME women with PCOS. In agreement, the Royal College of Obstetricians and Gynaecologists (RCOG) recommends the routine screening for psychological symptoms in all women with PCOS.^67^ Yet, our meta-analysis depicts that there is limited number of studies exploring psychological comorbidities in PCOS patients, both in UK and worldwide. Additionally, our meta-analysis was unable to meet most pre-planned outcomes, including the effects of ethnicity and race on MH sequelae in PCOS women, due to the lack of data identified within systematically pooled studies. This was largely due to the non-reporting of patient race and ethnicity in PCOS studies examining patient MH.

The two most identified MH conditions amongst PCOS patients in our systematic review were depression and anxiety. Indeed, several existing studies comment on the high risk of both existing and new diagnoses of depression in women with PCOS.^68,69^ Our meta-analysis confirmed these findings, as major depression affected 17% of PCOS patients, and elevated existing depression affected 33%, with a considerably high heterogeneity in both subthemes within and between regions. Existing literature in the field measuring true prevalence of depression in BAME patients with PCOS is contradictory. This meta-analysis could not pool race or ethnicity with the data due to differences in research study designs and clinical practices, as well as the lack of BAME patient representation within studies. Therefore, the true prevalence of elevated existing depression and newly diagnosed major depression amongst BAME PCOS women could not be estimated. However, our narrative synthesis identified certain races/ethnicities to carry higher rates of depression than Caucasian PCOS patients.

Anxiety disorders include generalised anxiety disorder, panic disorder, separation anxiety disorder, social anxiety disorder and specific phobias^70^ and may impair daily functioning. Women are twice as likely to develop anxiety disorders than men,^70^ with women with PCOS have a significantly higher prevalence of generalised anxiety symptoms than women without PCOS.^71^ This study revealed that there are a small number of published studies commenting on anxiety prevalence in women with PCOS. Of these, we identified 41% prevalence of generalised anxiety disorder among women with PCOS, however, heterogeneity was high at 94.83%. This heterogeneity extended to results from different regions suggests lack of difference between geographical locations in prevalence of anxiety amongst women with PCOS. Similar to our depression subtheme analysis, we were unable to establish the prevalence of anxiety amongst different ethnicities and races in women with PCOS due to lack of robust data.

Eating Disorders (EDs) have high prevalence particularly in young women worldwide.^72^ Body image disturbances and self-esteem are central to the development of common EDs and conditions that may affect body image has been found to contribute to the development of EDs.^73^ The body dissatisfaction associated with features such as weight gain, hyperandrogenism, hirsutism, acne, and androgenic alopecia often contribute to disturbed body image perception linked with EDs. Existing literature, although informative on the relationship between PCOS and EDs, struggles to report findings on ethnic differences in ED and PCOS. Only two observational studies were identified addressing EDs in BAME PCOS patients,^74,75^ and although they report an increased risk of disordered eating habits in PCOS women compared with the control populations, in agreement, one study found Black, Hispanic and Mixed race women with PCOS to have higher EDE-Q scores (a self-rating questionnaire of the range and severity of ED features) compared with White women^75^ yet the other reported EDs to be similar in the BAME PCOS population.^75^ However, discordant methodology and the use of external cohorts from a different country, makes the available evidence to be substandard to draw firm conclusions on the specific prevalence of EDs in BAME women with PCOS, indicating the need for further research to understanding ethnic differences in PCOS and associated EDs.

Psychotic disorders, such as schizophrenia have been reported to be significantly more prevalent in patients with PCOS in a national study in Sweden.^76^ However, there is a dearth of studies investigating the associations between conditions. Although people from BAME background had been reported to experience a higher prevalence rate of psychosis^77^ compared to the Caucasian population in UK, only one cohort study explored the prevalence of psychotic disorders in Taiwanese PCOS patients,^78^ and reported no statistical difference in prevalence of schizophrenia between the two populations.^78^ This highlights the need for future high quality studies to assess psychotic illnesses relevant to PCOS amongst different ethnicities.

Bipolar disorder (BD) has been reported to be more prevalent in women with PCOS^79^ compared to the general population and since it carries a 12-fold increased risk of suicide,^80^ it is especially important to examine any causative common mechanistic relations between the two conditions. However, there is a caveat, in that there is a reported high prevalence of menstrual disorders in women receiving treatment for BD.^81^ Therefore, accounting for confounding variables such as medication and pre-existing conditions is vital in studies that assess prevalence of MH sequalae, such as BD, in BAME PCOS women. A study from Kashmir Valley^82^ reported BD rates of 2.72% compared to 0.00% in control participants while a nationwide cohort study from Taiwan^78^ found no significant alteration in prevalence of BD in PCOS women compared to a control population. Further research is thus required in order to develop understanding of the prevalence in different populations, as associations cannot be drawn with the limited data available.

This systematic review had further revealed the heterogeneity of diagnosing psychiatric disorders. Clinicians have adopted 8 different self-report questionnaires in their methodologies highlighted by this review. This limits our study’s generalisability as well as any estimate of prevalence of a given psychiatric disorder.

Overall, although most studies used Rotterdam Criteria in the diagnosis of PCOS,^5^ 4 studies adopted the National Institutes of Health Criteria, 1990 to diagnose PCOS, which will bring obvious diagnostic bias.^83–86^ Future studies should strive to adapt internationally accepted, standardised criteria for the diagnosis of both PCOS and MH disorder to avoid sampling bias.

Medical practitioners may not be aware of the impact different cultures may have on patient’s reporting of both physical and psychiatric symptoms . In general, members of BAME communities are less likely to engage with MH services or seek help before symptoms severely impact their function.^87,88^ Simkhada et al. found that members of Nepali and Iranian communities report improved MH care and support by healthcare professionals who were trained on different cultural practices.^89^ Furthermore, clinicians in the study stated that a better understanding of different cultures to be beneficial for providing a culturally sensitive service. Therefore, training healthcare professionals in cultural norms within BAME communities is vital to better understand their knowledge and beliefs of MH symptoms, thereby allowing clinicians to provide a holistic, effective treatment approach when working with diverse populations. Cultural competency training may also increase patient trust in clinicians and lead to early discussions of how PCOS may be affecting patients psychosocially within distinct BAME communities. This will therefore aid in early detection and effective and lasting treatment of MH sequelae of PCOS and personalise patient clinical care.

## Limitations

Several limitations affected our systematic review and meta-analysis findings. Whilst studies listed often identified severe emotional distress amongst PCOS women, some of this was self-reported and therefore subjective, without the presence of a clinical diagnosis. This may mask the true prevalence of common MH conditions, such as depression and anxiety, amongst women with PCOS. A further limitation includes the high heterogeneity observed across all subtheme meta-analyses. Several sources of heterogeneity were behind the variations across estimates. These include the range of scales used, the choice of threshold, although many are equivalent but nonetheless, some disagreement remains and contributes to variations. Other factors such as differences in age range, body mass index (BMI), region, study design, sample size and methodology, population ethnicity and other charactersitics, and the established unobserved heterogeneity, all tapping into the differences observed across studies.

The cultural differences in reporting MH symptoms across different regions, potentially further impacted by complex interaction with stigma and accepted norms within BAME communities may have specific effects on different populations. Moreover, the difference in reporting MH symptoms and diagnoses is a further limitation of all studies included in this review. Several diverse assessment tools were used in the assessment of risk of key psychiatric disorders, including self-reported symptoms, self-reported use of psychiatric medication and clinical diagnosis, thereby introducing heterogeneity in studies included in this review. This may limit the generalisability of the data obtained, may mask the true prevalence of a given psychiatric disorder due to questionnaires’ tendency to screen rather than diagnose.

## Conclusion

This study demonstrates the lack of ethnic minority representation in research studies conducted among PCOS patients exploring a possible MH sequalae. To determine if there is a bi-directional relationship between PCOS and MH conditions among ethnic minorities, comprehensive research studies should be designed and conducted as part of a global initiative. A key attribute to the changing needs within women’s physical and MH is the migratory patterns that changes the regional and global population. The associated nuances influence both clinical practice and the access to the health-care system, and therefore, regular, and careful scrutiny of contemporary evidence is essential to optimise the overall clinical care offered to these patients. As a result, cultural appropriation based training should be made available and accessible to healthcare professionals within primary and secondary care settings.

## Data Availability

All data produced in the present study are available upon reasonable request to the authors

## Data sharing statement

All data used within this study has been publicly available. The authors will consider the sharing the dataset gathered upon request.

## Contributors

GD and DKH developed the systematic review protocol and embedded this within the ELEMI project’s evidence synthesis phase. GD, VB, DKH, SR, + SA wrote the first draft of the manuscript. This was furthered by PP, AS, WG, NT, YZ, JS and SR. KM, GD, YZ, VB and JS shared database searches, study selection and extraction for analysis. SA and GD conducted the analysis including the design of the statistical analysis plan. GD, YZ, DKH, PP, KE, WG, NT, YZ, JS, KM, VR, AS, PB, HM, AM, WG and KM critically appraised and finalised the manuscript. All authors approved the final version of the manuscript.

## Acknowledgements

The authors acknowledge support from Southern Health NHS Foundation Trust, and Liverpool Women’s hospital. We would like to acknowledge Mrs Nyla Haque who inspired the discussion of BAME groups within the context of this study.

This paper is part of the multifaceted ELEMI project that is sponsored by Southern Health NHS Foundation Trust and in collaboration with the University of Liverpool, University College London, University College London NHS Foundation Trust, Liverpool Women’s Hospital, Robinson Research Institute (University of Adelaide), Ramaiah Memorial Hospital (India), University of Geneva and Manchester University NHS Foundation Trust.

## Supplementary Materials

All supplementary materials associated with this article can be found in the attached Appendices.

---

**Supplementary a1:**
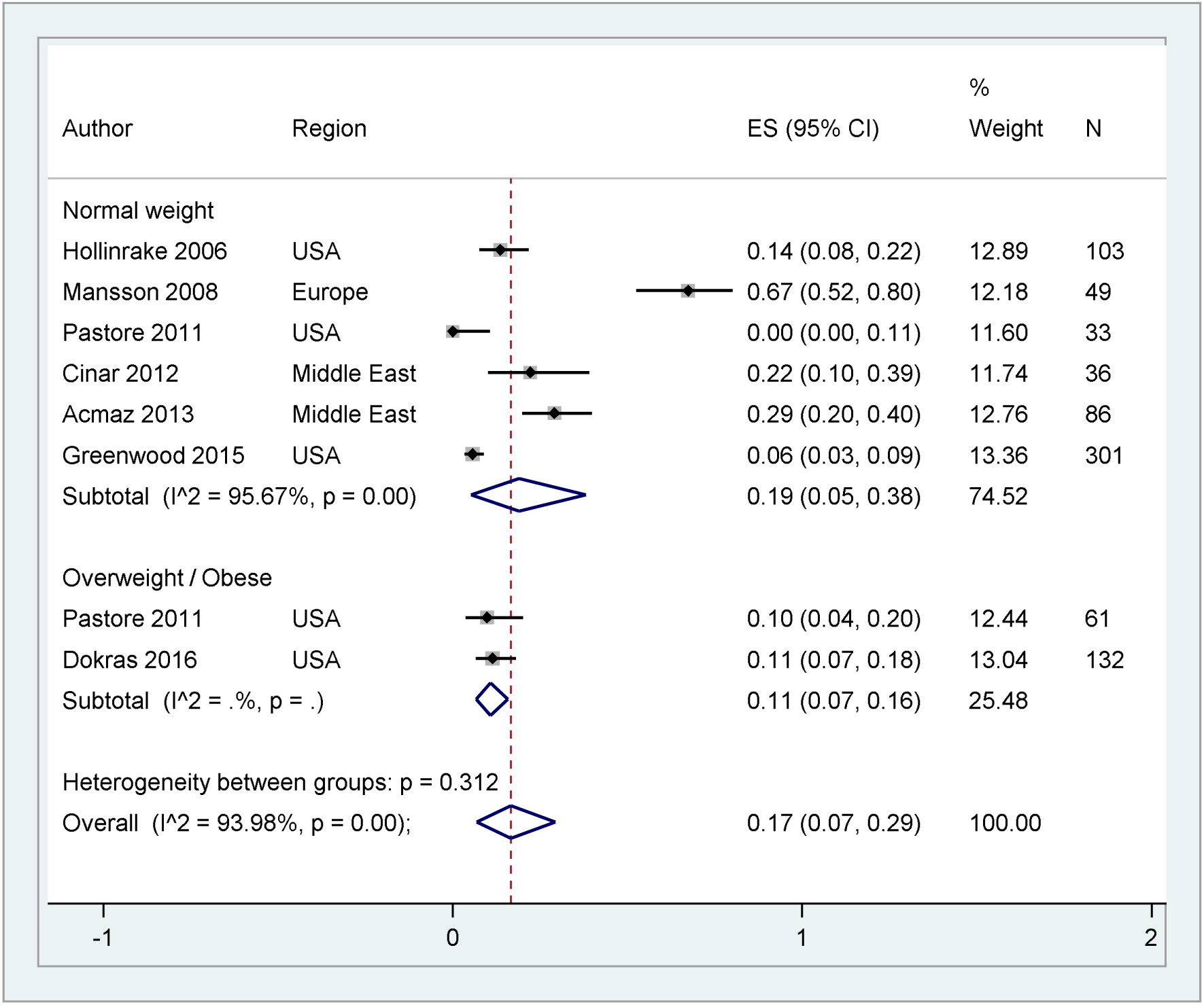
Major depression by BMI (sensitivity)

**Supplementary a2:**
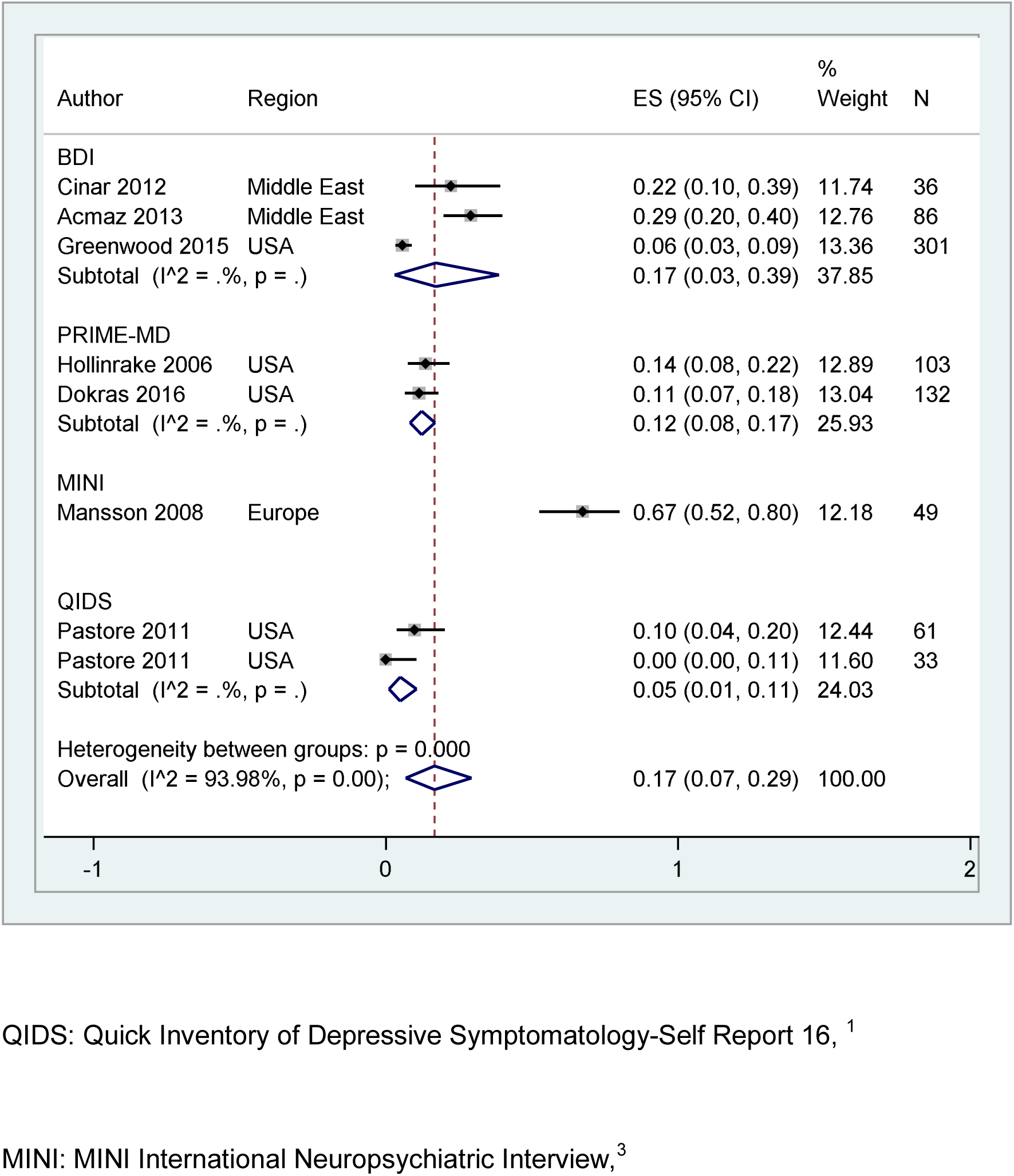
Major depression by assessment tool (sensitivity)

**Supplementary b1:**
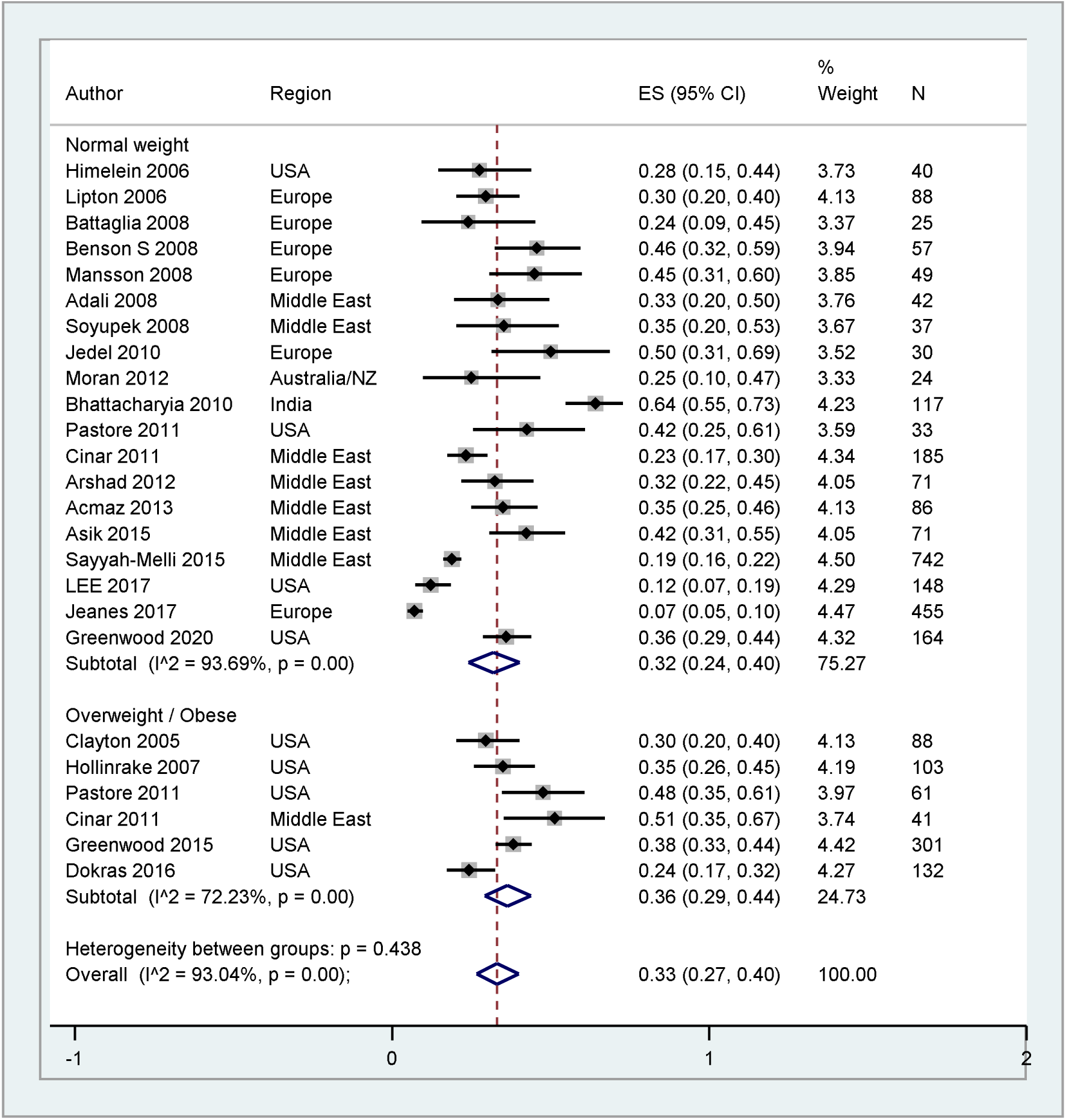
elevated depression by BMI (sensitivity)

**Supplementary b2:**
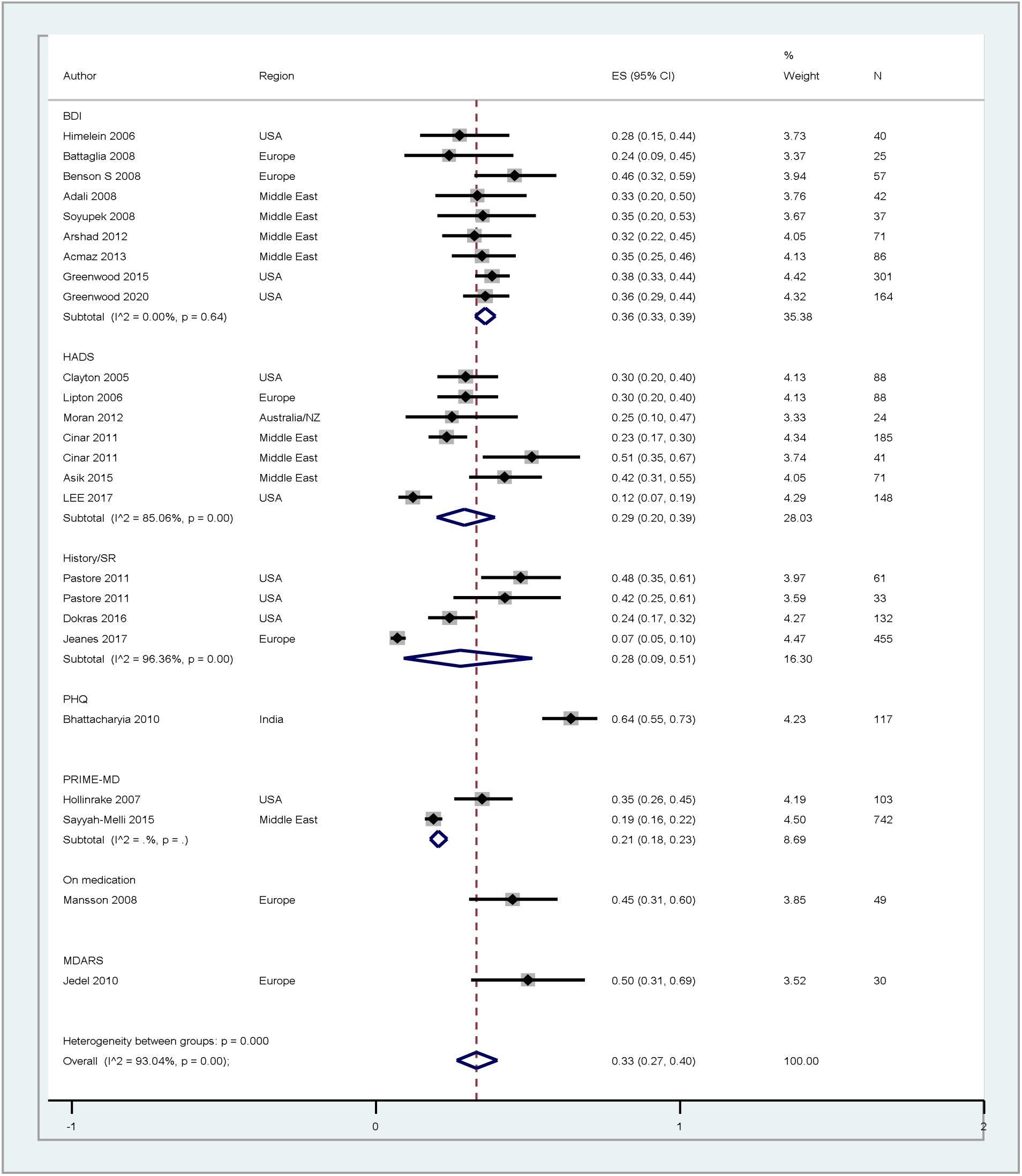
elevated depression by assessment tool (sensitivity)

**Supplementary c1:**
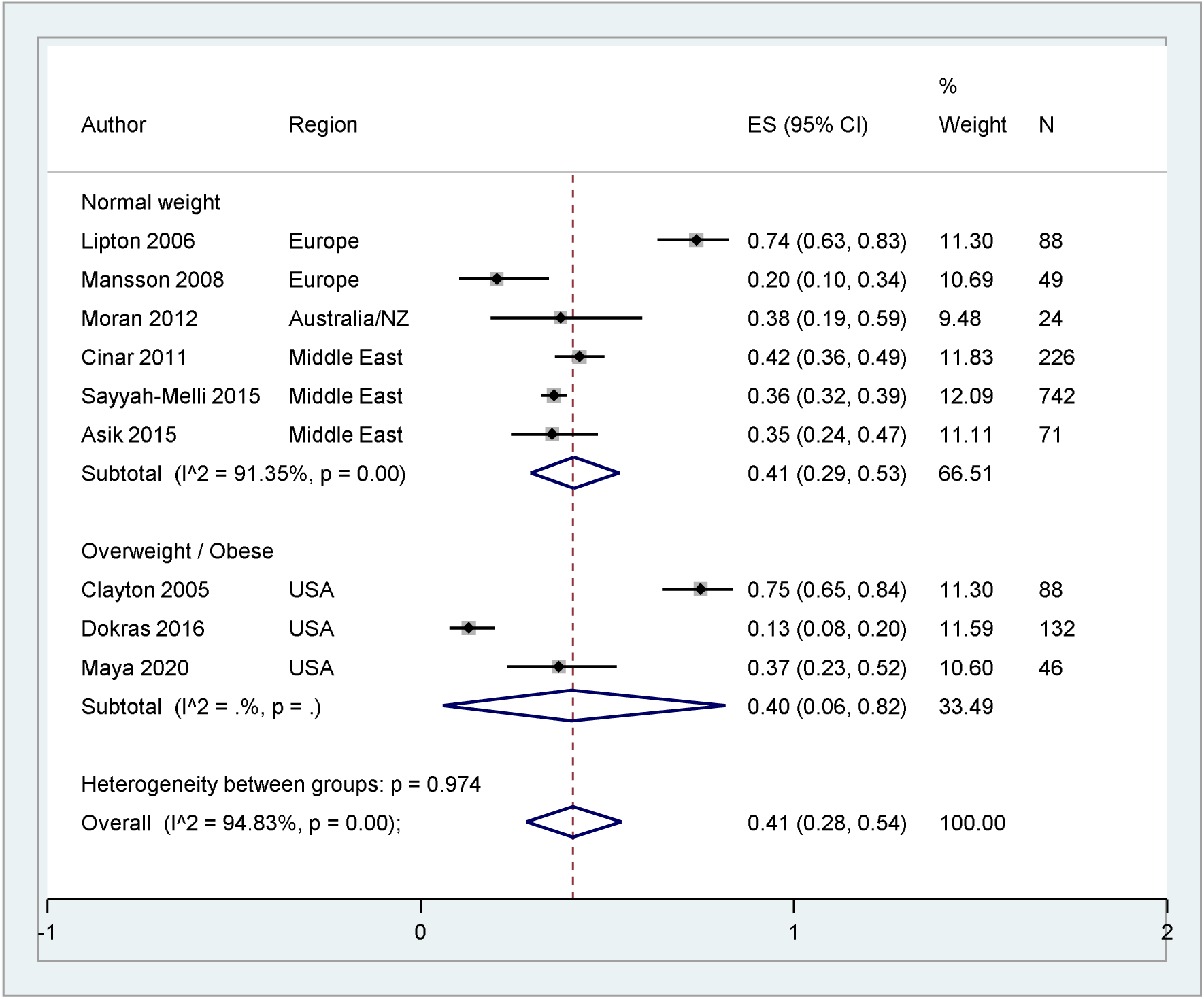
Prevalence of anxiety by BMI (sensitivity)

**Supplementary c2:**
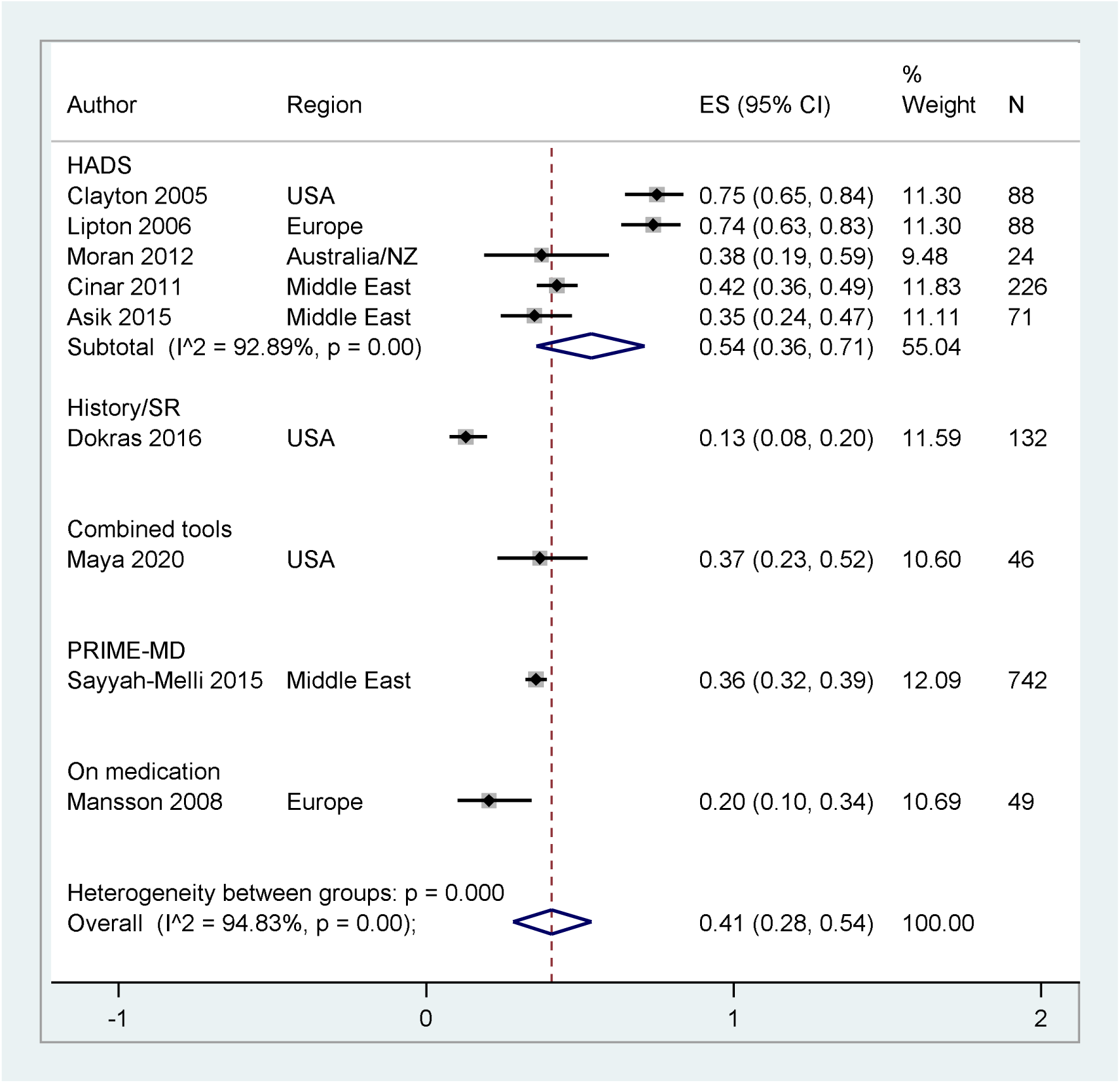
Prevalence of Anxiety by assessment tool (sensitivity)

**Supplementary d1:**
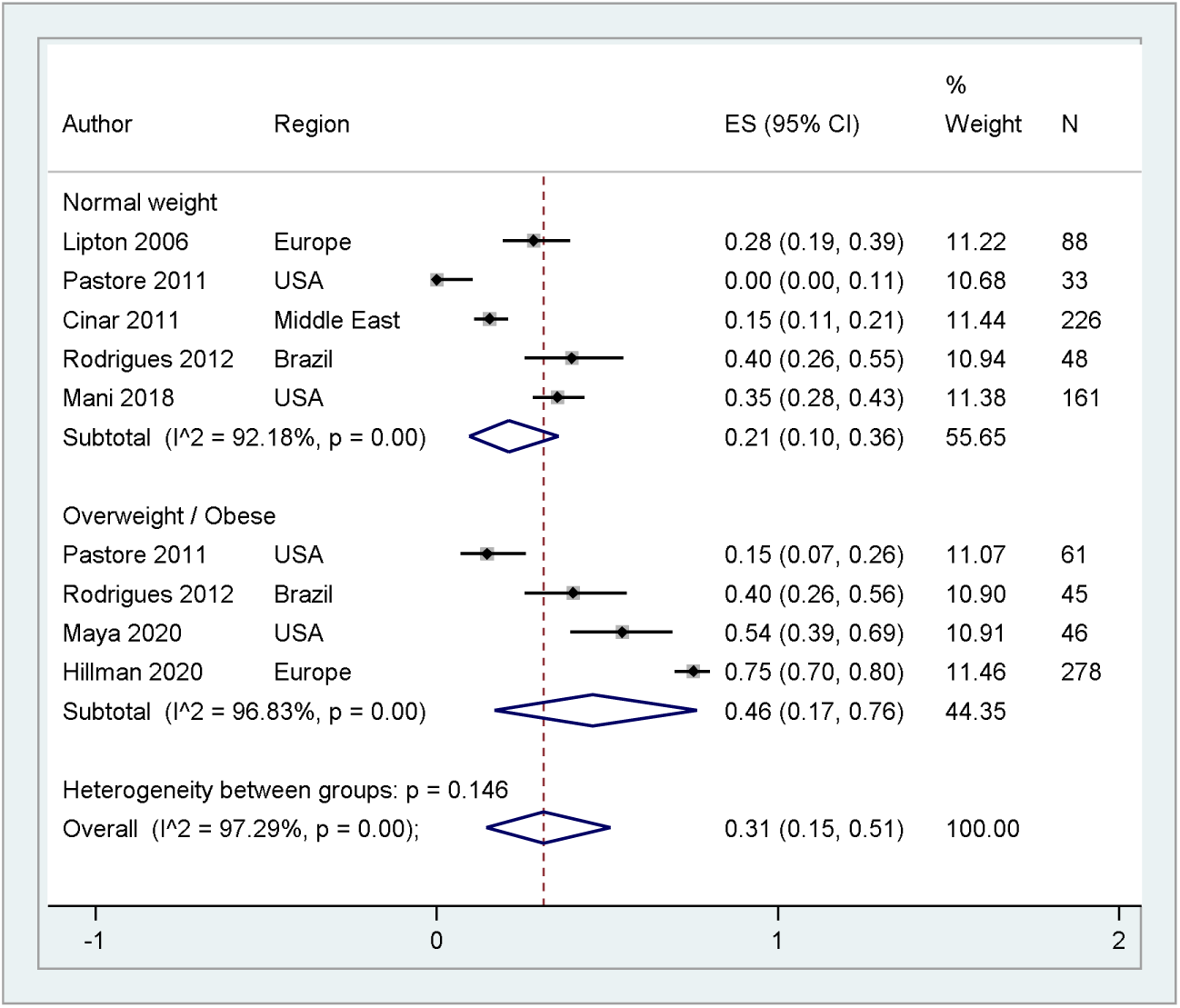
Common mental health by BMI (sensitivity)

**Supplementary d:**
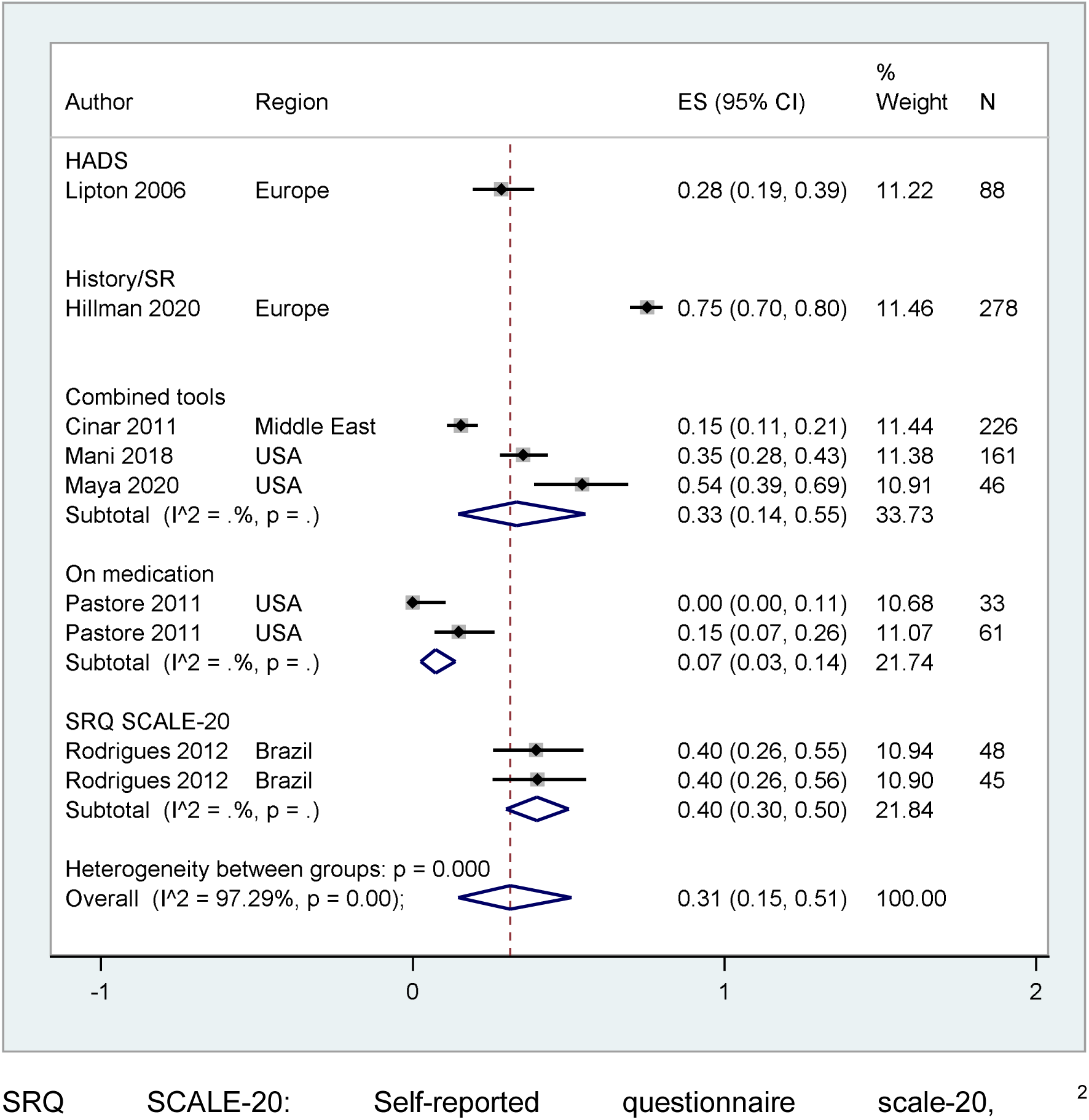
Common mental health by assessment tool (sensitivity)

## References

Abu Hashim H. Twenty years of ovulation induction with metformin for PCOS; what is the best available evidence?. Reprod Biomed Online. 2016;32(1):44–53. doi:10.1016/j.rbmo.2015.09.015

Bjonnes AC, Saxena R, Welt CK. Relationship between polycystic ovary syndrome and ancestry in European Americans. Fertil Steril. 2016;106(7):1772–1777. doi:10.1016/j.fertnstert.2016.08.033

Ding T, Hardiman PJ, Petersen I, Wang FF, Qu F, Baio G. The prevalence of polycystic ovary syndrome in reproductive-aged women of different ethnicity: a systematic review and meta-analysis. Oncotarget. 2017;8(56):96351–96358. doi:10.18632/oncotarget.19180

Gibson-Helm M, Teede H, Dunaif A, Dokras A. Delayed Diagnosis and a Lack of Information Associated With Dissatisfaction in Women With Polycystic Ovary Syndrome. J Clin Endocrinol Metab. 2017;102(2):604–612. doi:10.1210/jc.2016-2963

Greenhill C. Mechanism underlying beneficial effects of exercise in PCOS identified. Nat Rev Endocrinol. 2018;14(8):441. doi:10.1038/s41574-018-0041-1

Koch L. Androgens inversely related to depression in PCOS. Nat Rev Endocrinol. 2011;7:438. doi:10.1038/nrendo.2011.97

Tzalazidis R, Oinonen KA. Continuum of Symptoms in Polycystic Ovary Syndrome (PCOS): Links with Sexual Behavior and Unrestricted Sociosexuality. J Sex Res. 2020;1–13.

Yildizhan R, Gokce AI, Yildizhan B, Cim N. Comparison of the effects of chlormadinone acetate versus drospirenone containing oral contraceptives on metabolic and hormonal parameters in women with PCOS for a period of two-year follow-up. Gynecol Endocrinol. 2015;31(5):396–400. doi:10.3109/09513590.2015.1006187

The Lancet Diabetes Endocrinology. Empowering women with PCOS. Lancet Diabetes Endocrinol. 2019;7(10):737. doi:10.1016/S2213-8587(19)30289-X

## References

1. Azziz R, Carmina E, Dewailly D, Diamanti-Kandarakis E, Escobar-Morreale HF, Futterweit W. Position statement: criteria for defining polycystic ovary syndrome as a predominantly hyperandrogenic syndrome: an Androgen Excess Society guideline. J Clin Endocrinol Metab. 2006;91(11):4237–4245. doi:10.1210/jc.2006-0178

2. Diamanti-Kandarakis E, Kandarakis H, Legro RS. The role of genes and environment in the etiology of PCOS. Endocrine. 2006;30(1):19–26. doi:10.1385/ENDO:30:1:19

3. March WA, Moore VM, Willson KJ, Phillips DI, Norman RJ, Davies MJ. The prevalence of polycystic ovary syndrome in a community sample assessed under contrasting diagnostic criteria. Hum Reprod. 2010;25(2):544–551. doi:10.1093/humrep/dep399

4. Bozdag G, Mumusoglu S, Zengin D, Karabulut E, Yildiz BO. The prevalence and phenotypic features of polycystic ovary syndrome: a systematic review and meta-analysis. Human Reprod. 2016;31(12):2841–2855. doi:10.1093/humrep/dew218

5. Teede H, Misso M, Costello M, et al. International evidence-based guideline for the assessment and management of polycystic ovary syndrome (PCOS). European Society of Human Reproduction and Embryology. 2018. https://www.eshre.eu/Guidelines-and-Legal/Guidelines/Polycystic-Ovary-Syndrome

6. Wolf WM, Wattick RA, Kinkade ON, Olfert MD. Geographical Prevalence of Polycystic Ovary Syndrome as Determined by Region and Race/Ethnicity. Int J Environ Res Public Health. 2018;15(11):2589. doi:10.3390/ijerph15112589

7. The Rotterdam ESHRE/ASRM-Sponsored PCOS Consensus Workshop Group. Revised 2003 consensus on diagnostic criteria and long-term health risks related to polycystic ovary syndrome. Fertil Steril. 2004;81(1):19–25. doi:10.1016/j.fertnstert.2003.10.004

8. Sendur SN, Yildiz BO. Influence of ethnicity on different aspects of polycystic ovary syndrome: a systematic review. Reprod BioMed Online. 2021;42(4):799–818. doi: 10.1016/j.rbmo.2020.12.006

9. Engmann L, Jin S, Sun F, et al. Racial and ethnic differences in the polycystic ovary syndrome metabolic phenotype. Am J Obstet Gynecol. 2017;216(5):493.e1-493.e13. doi:10.1016/j.ajog.2017.01.003

10. Escobar-Morreale HF. Polycystic ovary syndrome: definition, aetiology, diagnosis and treatment. Nat Rev Endocrinol. 2018;14(5):270–284. doi:10.1038/nrendo.2018.24

11. Amato P, Simpson JL. The genetics of polycystic ovary syndrome. Best Pract Res Clin Obstet Gynaecol. 2004;18(5):707–718. doi:10.1016/j.bpobgyn.2004.05.002

12. Azziz R, Carmina E, Chen Z, et al. Polycystic ovary syndrome. Nat Rev Dis Primers. 2016;2:16057. doi:10.1038/nrdp.2016.57

13. Apridonidze T, Essah PA, Iuorno MJ, Nestler JE. Prevalence and characteristics of the metabolic syndrome in women with polycystic ovary syndrome. J Clin Endocrinol Metab. 2005;90(4):1929–1935. doi:10.1210/jc.2004-1045

14. Legro RS, Kunselman AR, Dodson WC, Dunaif A. Prevalence and predictors of risk for type 2 diabetes mellitus and impaired glucose tolerance in polycystic ovary syndrome: a prospective, controlled study in 254 affected women. J Clin Endocrinol Metab. 1999;84(1):165–169. doi:10.1210/jcem.84.1.5393

15. Teede H, Deeks A, Moran L. Polycystic ovary syndrome: a complex condition with psychological, reproductive and metabolic manifestations that impacts on health across the lifespan. BMC Medicine. 2010;8(41):1–10. doi:10.1186/1741-7015-8-41

16. Moran L, Gibson-Helm M, Teede H, Deeks A. Polycystic ovary syndrome: a biopsychosocial understanding in young women to improve knowledge and treatment options. J Psychosom Obstet Gynecol. 2010;31(1):24–31. doi:10.3109/01674820903477593

17. Hillman SC, Bryce C, Caleyachetty R, Dale J. Women’s experiences of diagnosis and management of polycystic ovary syndrome: a mixed-methods study in general practice. Br J Gen Pract. 2020;70(694):e322–e329. doi:10.3399/bjgp20X708881

18. Dokras A, Stener-Victorin E, Yildiz BO, et al. Androgen excess — Polycystic Ovary Syndrome Society: position statement on depression, anxiety, quality of life, and eating disorders in polycystic ovary syndrome. Fertil Steril. 2018;109(5):888–899. doi:10.1016/j.fertnstert.2018.01.038

19. Balen A. Polycystic ovary syndrome. InnovAiT. 2016;9(3):144–150.

20. Kumarendran B, Sumilo D, O’Reilly MW, et al. Increased risk of obstructive sleep apnoea in women with polycystic ovary syndrome: a population-based cohort study. Eur J Endocrinol. 2019;180(4):265–272. doi:10.1530/EJE-18-0693

21. Dokras A. Mood and anxiety disorders in women with PCOS. Steroids. 2012;77(4):338–341. doi:10.1016/j.steroids.2011.12.008

22. Davari-Tanha F, Hosseini Rashidi B, Ghajarzadeh M, Noorbala AA. Bipolar disorder in women with polycystic ovarian syndrome (PCO). Acta Med Iran. 2014;52(1):46–48.

23. Kerchner A, Lester W, Stuart SP, Dokras A. Risk of depression and other mental health disorders in women with polycystic ovary syndrome: a longitudinal study. Fertil Steril. 2009;91(1):207–212. doi:10.1016/j.fertnstert.2007.11.022

24. Farrell K, Antoni MH. Insulin resistance, obesity, inflammation, and depression in polycystic ovary syndrome: biobehavioral mechanisms and interventions. Fertil Steril. 2010;94(5):1565–1574. doi:10.1016/j.fertnstert.2010.03.081

25. Hollinrake E, Abreu A, Maifeld M, Van Voorhis BJ, Dokras A. Increased risk of depressive disorders in women with polycystic ovary syndrome. Fertil Steril. 2007;87(6):1369–1376. doi:10.1016/j.fertnstert.2006.11.039

26. Yin X, Ji Y, Chan CLW, Chan CHY. The mental health of women with polycystic ovary syndrome: a systematic review and meta-analysis. Arch Womens Ment Health. 2021;24(1):11–27. doi:10.1007/s00737-020-01043-x

27. Prince M, Patel V, Saxena S, et al. No health without mental health. Lancet. 2007;370(9590):859–877. doi:10.1016/S0140-6736(07)61238-0

28. Memon A, Taylor K, Mohebati LM, et al. Perceived barriers to accessing mental health services among black and minority ethnic (BME) communities: a qualitative study in Southeast England. BMJ Open. 2016;6:e012337. doi:10.1136/bmjopen-2016-012337

29. American Psychiatric Association. Diagnostic and statistical manual of mental disorders. 5th ed. 2013:21.

30. Spitzer RL, Kroenke K, Williams JB. Validation and utility of a self-report version of PRIME-MD: the PHQ primary care study. Primary Care Evaluation of Mental Disorders. Patient Health Questionnaire. JAMA. 1999;282(18):1737–1744. doi:10.1001/jama.282.18.1737

31. Snaith RP, Zigmond AS. The hospital anxiety and depression scale. Br Med J (Clin Res Ed). 1986;292(6516):344. doi:10.1136/bmj.292.6516.344

32. Beck AT, Steer RA, Brown GK. Beck depression inventory (BDI-II). Pearson; 1996.

33. Davidson J, Turnbull CD, Strickland R, et al. The Montgomery - Åsberg Depression Scale: reliability and validity. Acta Psychiatr Scand. 1986;73(5):544–548. doi:10.1111/j.1600-0447.1986.tb02723.x

34. Devins GM, Orme CM, Costello CG, et al. Measuring depressive symptoms in illness populations: Psychometric properties of the Center for Epidemiologic Studies Depression (CES-D) scale. Psychology and Health. 1988;2(2):139–156. doi:10.1080/08870448808400349

35. Score IRC. The Reynolds Adolescent Depression Scale-(RADS-2). Comprehensive Handbook of Psychological Assessment, Volume 2: Personality Assessment. 2003;2:224.

36. REFERENCE MISSING

37. REFERENCE MISSING

38. Spitzer RL, Kroenke K, Williams JB, et al. A brief measure for assessing generalized anxiety disorder: the GAD-7. Arch Interl Med. 2006;166(10):1092–1097. doi:10.1001/archinte.166.10.1092

39. REFERENCE MISSING

40. Cinar N, Harmanci A, Demir B, et al. Effect of an oral contraceptive on emotional distress, anxiety and depression of women with polycystic ovary syndrome: a prospective study. Hum Reprod. 2012;27(6):1840–1845. doi:10.1093/humrep/des113

41. Benson J, Severn C, Hudnut-Beumler J, et al. Depression in Girls With Obesity and Polycystic Ovary Syndrome and/or Type 2 Diabetes. Can J Diabetes. 2020;44(6):507–513. doi:10.1016/j.jcjd.2020.05.015

42. Pastore LM, Patrie JT, Morris WL, et al. Depression symptoms and body dissatisfaction association among polycystic ovary syndrome women. J Psychosom Res. 2011;71(4):270–276. doi:10.1016/j.jpsychores.2011.02.005

43. Maya J, Siegel J, Cheng TQ, et al. Prevalence and risk factors of polycystic ovarian syndrome among an ethnically diverse overweight/obese adolescent population. Int J Adolesc Med Health. 2020. doi:10.1515/ijamh-2019-0109

44. Rodrigues CE, Ferreira Lde L, Jansen K, et al. Evaluation of common mental disorders in women with polycystic ovary syndrome and its relationship with body mass index. Rev Bras Ginecol Obstet. 2012;34(10):442–446. doi:10.1590/s0100-72032012001000002

45. DerSimonian R, Laird N. Meta-analysis in clinical trials. Controlled clinical trials. 1986;7(3):177–188. doi:10.1016/0197-2456(86)90046-2

46. Newcombe RG. Interval estimation for the difference between independent proportions: comparison of eleven methods. Statistics in medicine. 1998;17(8):873–890. doi:10.1002/(sici)1097-0258(19980430)17:8<873::aid-sim779>3.0.co;2-i

47. Newcombe RG. Two - sided confidence intervals for the single proportion: comparison of seven methods. Statistics in medicine. 1998;17(8):857–872. doi:10.1002/(sici)1097-0258(19980430)17:8<857::aid-sim777>3.0.co;2-e

48. Higgins JP, Thompson SG, Deeks JJ, et al. Measuring inconsistency in meta-analyses. Bmj 2003;327(7414):557–560. doi:10.1136/bmj.327.7414.557

49. REFERENCE MISSING

50. REFERENCE MISSING

51. REFERENCE MISSING

52. REFERENCE MISSING

53. REFERENCE MISSING

54. REFERENCE MISSING

55. REFERENCE MISSING

56. Hoeger KM, Dokras A, Piltonen T. Update on PCOS: Consequences, Challenges, and Guiding Treatment. J Clin Endocrinol Metab. 2021;106(3):e1071–e1083. doi:10.1210/clinem/dgaa839

57. Witchel SF, Teede HJ, Peña AS. Curtailing PCOS. Pediatr Res. 2020;87(2):353–361. doi:10.1038/s41390-019-0615-1

58. Lin AW, Bergomi EJ, Dollahite JS, Sobal J, Hoeger KM, Lujan ME. Trust in Physicians and Medical Experience Beliefs Differ Between Women With and Without Polycystic Ovary Syndrome. J Endocr Soc. 2018;2(9):1001–1009. doi:10.1210/js.2018-00181

59. Hillman SC, Bryce C, Caleyachetty R, Dale J. Women’s experiences of diagnosis and management of polycystic ovary syndrome: a mixed-methods study in general practice. Br J Gen Pract. 2020;70(694):e322–e329. doi:10.3399/bjgp20X708881

60. Saidunnisa BG, Shariff A, Ayman G, Mohammad B, Housam R, Khaled N. Assessment of Risk Factors for development of Polycystic Ovarian Syndrome. Int J Contemp Med Research. 2017;4(1):164–167.

61. Lo JC, Feigenbaum SL, Yang J, Pressman AR, Selby JV, Go AS. Epidemiology and Adverse Cardiovascular Risk Profile of Diagnosed Polycystic Ovary Syndrome. J Clin Endocrinol Metab. 2006;91(4):1357–1363. doi:10.1210/jc.2005-2430

62. Cooney LG, Dokras A. (2018) Beyond fertility: polycystic ovary syndrome and long-term health. Fertil Steril. 2018;110(5):794–809. doi:10.1016/j.fertnstert.2018.08.021

63. Azziz R, Woods KS, Reyna R, Key TJ, Knochenhauer ES, Yildiz BO. The prevalence and features of the polycystic ovary syndrome in an unselected population. J Clin Endocrinol Metab. 2004;89(6):2745–2749. doi:10.1210/jc.2003-032046

64. McManus S, Bebbington P, Jenkins R, Brugha T, eds. Mental health and wellbeing in England: Adult Psychiatric Morbidity Survey 2014. NHS Digital; 2016.

65. Gajwani R, Parsons H, Birchwood M, Singh SP. Ethnicity and detention: are Black and minority ethnic (BME) groups disproportionately detained under the Mental Health Act 2007?. Soc Psychiatry Psychiatr Epidemiol. 2016;51(5):703–711. doi:10.1007/s00127-016-1181-z

66. Linney C, Ye S, Redwood S, et al. “Crazy person is crazy person. It doesn’t differentiate”: an exploration into Somali views of mental health and access to healthcare in an established UK Somali community. Int J Equity Health. 2020;19(1):190. doi:10.1186/s12939-020-01295-0

67. Ledger WL, Atkin SL, Sathyapalan T. Long-term Consequences of Polycystic Ovary Syndrome: Green-top Guideline No. 33. 3rd ed. Royal College of Obstetricians and Gynaecologists; 2014. https://www.rcog.org.uk/globalassets/documents/guidelines/gtg_33.pdf

68. Greenwood EA, Yaffe K, Wellons MF, Cedars MI, Huddleston HG. Depression over the lifespan in a population-based cohort of women with polycystic ovary syndrome: longitudinal analysis. J Clin Endocrinol Metab. 2019;104(7):2809–2819. doi:10.1210/jc.2019-00234

69. Cooney LG, Dokras A. Depression and Anxiety in Polycystic Ovary Syndrome: Etiology and Treatment. Curr Psychiatry Rep. 2017;19(11):83. doi:10.1007/s11920-017-0834-2

70. Craske MG, Stein MB. Anxiety. Lancet. 2016;388(10063):3048–3059. doi:10.1016/S0140-6736(16)30381-6

71. REFERENCE MISSING

72. Galmiche M, Déchelotte P, Lambert G, Tavolacci MP. Prevalence of eating disorders over the 2000-2018 period: a systematic literature review. Am J Clin Nutr. 2019;109(5):1402–1413. doi:10.1093/ajcn/nqy342

73. Tay CT, Teede HJ, Hill B, Loxton D, Joham AE. Increased prevalence of eating disorders, low self-esteem, and psychological distress in women with polycystic ovary syndrome: a community-based cohort study. Fertil Steril. 2019;112(2):353–361. doi:10.1016/j.fertnstert.2019.03.027

74. Greenwood EA, Pasch LA, Cedars MI, Huddleston HG. Obesity and depression are risk factors for future eating disorder-related attitudes and behaviors in women with polycystic ovary syndrome. Fertil Steril. 2020;113(5):1039–1049. doi:10.1016/j.fertnstert.2020.01.016

75. Lee I, Cooney LG, Saini S, et al. Increased risk of disordered eating in polycystic ovary syndrome. Fertil Steril. 2017;107(3):796–802. doi:10.1016/j.fertnstert.2016.12.014

76. Cesta CE, Månsson M, Palm C, Lichtenstein P, Iliadou AN, Landén M. Polycystic ovary syndrome and psychiatric disorders: Co-morbidity and heritability in a nationwide Swedish cohort. Psychoneuroendocrinology. 2016;73:196–203. doi:10.1016/j.psyneuen.2016.08.005

77. Qassem T, Bebbington P, Spiers N, McManus S, Jenkins R, Dein S. Prevalence of psychosis in black ethnic minorities in Britain: analysis based on three national surveys. Soc Psychiatry Psychiatr Epidemiol. 2015;50(7):1057–1064. doi:10.1007/s00127-014-0960-7

78. Hung JH, Hu LY, Tsai SJ, et al. Risk of psychiatric disorders following polycystic ovary syndrome: a nationwide population-based cohort study. PLoS One. 2014;9(5):e97041. doi:10.1371/journal.pone.0097041

79. Brutocao C, Zaiem F, Alsawas M, Morrow AS, Murad MH, Javed A. Psychiatric disorders in women with polycystic ovary syndrome: a systematic review and meta-analysis. Endocrine. 2018;62(2):318–325. doi:10.1007/s12020-018-1692-3

80. Grunze H. Chapter 40 - Bipolar Disorder. In: Zigmond MJ, Rowland LP, Coyle JT, eds. Neurobiology of Brain Disorders. Academic Press, San Diego; 2015:655–673.

81. Rasgon NL, Altshuler LL, Fairbanks L, et al. Reproductive function and risk for PCOS in women treated for bipolar disorder. Bipolar Disord. 2005;7(3):246–259. doi:10.1111/j.1399-5618.2005.00201.x

82. Hussain A, Chandel RK, Ganie MA, et al. Prevalence of psychiatric disorders in patients with a diagnosis of polycystic ovary syndrome in kashmir. Indian J Psychol Med. 2015;37(1):66–70. doi:10.4103/0253-7176.150822

83. REFERENCE MISSING

84. REFERENCE MISSING

85. REFERENCE MISSING

86. REFERENCE MISSING

87. Casey, M. Health Needs Assessment of the Nepali Community in Rushmoor. Hampshire County Council. Updated October, 2010. https://documents.hants.gov.uk/public-health/NepaliHealthNeedsAssessment2010.pdf

88. Cooper C, Spiers N, Livingston G, et al. Ethnic inequalities in the use of health services for common mental disorders in England. Soc Psychiatry Psychiatr Epidemiol. 2013;48(5):685–692. doi:10.1007/s00127-012-0565-y

89. Simkhada B, Vahdaninia M, van Teijlingen E, Blunt H. Cultural issues on accessing mental health services in Nepali and Iranian migrants communities in the UK. Int J Ment Health Nurs. 2021. doi:10.1111/inm.12913

